# Identifying and Characterizing Gallstone Disease from Clinical Narratives with Zero-shot Learning and Automated Prompt Optimization

**DOI:** 10.64898/2026.01.29.26345132

**Authors:** Sy Hwang, Agnes Wang, Sunil Thomas, Ashley Batugo, David E Kaplan, Daniel J Rader, Danielle Mowery, Junghyun Lim

## Abstract

We built and evaluated a zero-shot LLM pipeline with automated, task-aware prompt optimization to extract radiology and symptom fields for gallstone phenotyping from de-identified EHR text. Across symptomatic, asymptomatic, and control cohorts, it performed reliably on high-signal binary fields and symptom flags but lagged on fine-grained stone burden and complications, establishing a practical baseline and motivating targeted refinements

## Introduction

Gallstone disease is highly prevalent and a cause of significant morbidity in the United States.^1^ Discernment of true cases and true controls is often challenging due to inconsistencies in translation of asymptomatic and symptomatic gallstone disease to International Classification of Diseases (ICD)-9/10 codes.^2^ In clinical practice, asymptomatic gallstones are often found incidentally on imaging (e.g. ultrasonography [US] and computed tomography [CT]) and are often overlooked/ not entered in as ICD-9/10 billing codes. Thus, relying solely on ICD-9/10 codes may lead to incorrectly assigning asymptomatic gallstone cases to controls. The need to reduce these inaccuracies emphasizes the need for the utilization of structured and unstructured data to identify gallstone cases. To our knowledge, no prior studies have published validation in the Epic Clarity regarding phenotype algorithms developed for gallstone disease via utilization of large language modeling (LLM).

## Methods

### Data

We developed a phenotype algorithm for gallstone disease based on structured and unstructured data incorporating an LLM algorithm, leveraging the Epic Clarity. We constructed a gallstone disease data mart of putative cases. Inclusion criteria consisted of adults (18 years old) with at least one abdominal imaging (US, CT, or magnetic resonance imaging [MRI]) from 1/1/2024 to 1/1/2025. Based on ICD and CPT codes, we categorized these individuals into symptomatic gallstone disease group (SGD), asymptomatic gallstone disease group (AGD), or no gallstone disease (NGD) based on the following criteria. For SGD, those with ICD codes for cholelithiasis, choledocholithiasis, cholecystitis, and gallstone pancreatitis, and CPT code for cholecystectomy and endoscopic retrograde cholangiopancreatography (ERCP) entered after the date of imaging were included. For AGD, those with ICD codes for cholelithiasis and choledocholithiasis entered after the date of imaging were included, and those with ICD codes for cholecystitis or gallstone pancreatitis (which reflect SGD) and CPT code for cholecystectomy or ERCP were excluded. For NGD, we excluded ICD codes for cholelithiasis, choledocholithiasis, cholecystitis, and gallstone pancreatitis, and CPT code for cholecystectomy and endoscopic retrograde cholangiopancreatography. For all individuals across the groups, we excluded individuals with ICD codes for acalculous cholecystitis, gallbladder polyps, biliary malignancy, pancreatic malignancy, neoplasm of ampulla of Vater, and sickle cell disease. We additionally excluded individuals with CPT codes for Whipple or liver transplant surgery and ERCP with non-gallstone-related procedures.

Upon categorizing the individuals into the three groups, we randomly sampled 100 patients from SGD, 50 patients from AGD, and 50 patients from NGD. We then collected structured data including demographics (age, race, sex), ICD and CPT codes. We also collected unstructured data which were free text from de-identified clinical notes, radiology reports, and pathology reports. Specifically, clinical notes (1 year back from the latest procedure date for SGD, from the latest abdominal imaging from AGD and NGD), radiology reports of the abdominal imaging from 1/1/2024 to 1/1/2025, and pathology reports from cholecystectomy that occurred following abdominal imaging. Among the randomly sampled patients, 100 SGD patients, 47 AGD patients, and 49 NGD patients had the notes and reports available for collection. All extracted variables were stored and analyzed in our HIPAA-compliant, Penn Medicine-managed server and project workspace. For these patients, a clinician with training in gastroenterology performed manual chart review, extracted data pertaining to symptoms, radiological evidence, and pathology descriptions of gallstone disease, and verified SGD, AGD, and NGD groups. Upon chart review, 118 patients were verified as SGD, 26 patients as AGD, and 52 patients as NGD.

### Prompting

We framed the study as a zero-shot instruction task. For each patient, the LLM was provided with patient’s de-identified notes within the study window, ordered chronologically. The prompt instructed the model to base all judgments only on the supplied corpus. For radiology, the model was tasked to capture to presence or absence of gallstones, gallbladder and gallbladder sludge. The model was instructed to extract potential complications including cholecystitis and extrahepatic duct dilation, as well as stone burden. For symptoms, the model was tasked with recording an overall symptom flag with structured pain attributes like location, radiation, character, onset, frequency and relation to food.

To improve instruction quality and task adherence, we perform task-aware automated prompt optimization ^**3**^, which involves two iterative processes. First, the system generates multiple candidate instruction prompts based on carefully curated initial prompts, producing variations in style, specificity and phrasing. Second, a critic phase scores each candidate prompt and provides targeted feedback based on classification performance. This process involves lightweight reasoning steps to guide consistent mapping from text spans to categorization through chain-of-thought. Candidates are then re-scored according to a selected task metric and the best instruction prompts are retained. This process iterates until either performance plateaus or a budget maximum is reached, whichever comes first, and yields a final prompt that is faithful to constraints and robust to variability in notes.

For model selection, we evaluated several instruction-tuned models and chose OpenAI’s GPT-4.1 as it delivered the best balance of latency, cost and label fidelity for our classification tasks. It produced well-formed outputs with fewer normalization corrections required post-hoc and had sufficient context size for the amount of input text we needed to provide. We set the sampling temperature to 0.1 to minimize stochastic variability, which proved to be critical when our outputs had to contain strictly enumerated label values. This configuration gave us adequate sensitivity while reducing spurious misclassification.

## Results

The model was reliable on high-signal binary fields (presence of gallbladder F1≈0.90–0.93; presence of gallstones F1≈0.70–0.77) and symptom presence when clearly documented (F1≈0.62–0.84 across cohorts), but it underperformed on fine-grained attributes such as stone count, size, and location (F1≈0.44–0.59) and on complications like cholecystitis, particularly in the asymptomatic cohort (F1≈0.40–0.62) (See Appendix). Error patterns reflect narrative heterogeneity and underspecification. Conservative defaults (“No/NA”) likely improved precision at the expense of recall. This presents a serviceable baseline and points to clear avenues for improvement.

## Discussion and Conclusions

Zero-shot, schema-constrained prompting produced a workable baseline for gallstone phenotyping with minimal engineering. It is sufficient for coarse cohort characterization and triage but remains limited on fine-grained attributes and complications due to narrative variability and temporal ambiguity. Next steps include post-hoc deterministic checks, developing proxy labels, few-shot exemplars, retrieval augmentation and fine-tuning.

## Data Availability

All data produced by the study is not openly available due to sensitivity of the dataset.

## Appendix

|  | Accuracy | Precision | Recall | F1 |
| --- | --- | --- | --- | --- |
| **Bucket 1 — Symptomatic Gallstone Disease** |  |  |  |  |
| **SYMPTOMS** |  |  |  |  |
| symptom_presence | 0.80 | 0.79 | 0.78 | 0.78 |
| abdominal_pain_presence | 0.78 | 0.77 | 0.76 | 0.76 |
| pain_location | 0.64 | 0.63 | 0.61 | 0.62 |
| pain_radiation | 0.58 | 0.56 | 0.55 | 0.55 |
| pain_character | 0.57 | 0.54 | 0.52 | 0.53 |
| pain_onset | 0.55 | 0.51 | 0.49 | 0.50 |
| pain_frequency | 0.60 | 0.58 | 0.56 | 0.57 |
| relation_to_food | 0.61 | 0.60 | 0.57 | 0.58 |
| nausea_vomiting | 0.64 | 0.63 | 0.58 | 0.60 |
| **RADIOLOGY** |  |  |  |  |
| presence_of_gallstone | 0.76 | 0.75 | 0.73 | 0.74 |
| presence_of_gallbladder | 0.91 | 0.90 | 0.90 | 0.90 |
| number_of_stones | 0.59 | 0.53 | 0.51 | 0.52 |
| diameter_of_largest_stone | 0.55 | 0.50 | 0.47 | 0.48 |
| location_of_stone | 0.62 | 0.60 | 0.58 | 0.59 |
| presence_of_gallbladder_sludge | 0.59 | 0.57 | 0.53 | 0.55 |
| cholecystitis | 0.65 | 0.64 | 0.60 | 0.62 |
| intrahepatic_duct_dilation | 0.60 | 0.58 | 0.55 | 0.57 |
| extrahepatic_duct_dilation | 0.59 | 0.57 | 0.54 | 0.56 |
| **Bucket 2 — Asymptomatic Gallstone Disease** |  |  |  |  |
| **SYMPTOMS** |  |  |  |  |
| symptom_presence | 0.66 | 0.63 | 0.61 | 0.62 |
| abdominal_pain_presence | 0.64 | 0.60 | 0.57 | 0.58 |
| pain_location | 0.56 | 0.52 | 0.49 | 0.50 |
| pain_radiation | 0.55 | 0.51 | 0.49 | 0.50 |
| pain_character | 0.53 | 0.49 | 0.46 | 0.47 |
| pain_onset | 0.52 | 0.47 | 0.42 | 0.44 |
| pain_frequency | 0.54 | 0.50 | 0.47 | 0.48 |
| relation_to_food | 0.56 | 0.53 | 0.49 | 0.51 |
| nausea_vomiting | 0.57 | 0.54 | 0.51 | 0.52 |
| **RADIOLOGY** |  |  |  |  |
| presence_of_gallstone | 0.78 | 0.78 | 0.76 | 0.77 |
| presence_of_gallbladder | 0.92 | 0.91 | 0.91 | 0.91 |
| number_of_stones | 0.57 | 0.52 | 0.48 | 0.50 |
| diameter_of_largest_stone | 0.54 | 0.49 | 0.42 | 0.44 |
| location_of_stone | 0.58 | 0.56 | 0.53 | 0.55 |
| presence_of_gallbladder_sludge | 0.56 | 0.52 | 0.49 | 0.51 |
| cholecystitis | 0.52 | 0.46 | 0.37 | 0.40 |
| intrahepatic_duct_dilation | 0.53 | 0.49 | 0.41 | 0.45 |
| extrahepatic_duct_dilation | 0.54 | 0.50 | 0.43 | 0.46 |
| **Bucket 3 — No Symptomatic Gallstone Disease** |  |  |  |  |
| **SYMPTOMS** |  |  |  |  |
| symptom_presence | 0.86 | 0.84 | 0.83 | 0.84 |
| abdominal_pain_presence | 0.84 | 0.82 | 0.80 | 0.81 |
| pain_location | 0.62 | 0.58 | 0.54 | 0.56 |
| pain_radiation | 0.62 | 0.60 | 0.56 | 0.58 |
| pain_character | 0.59 | 0.56 | 0.52 | 0.54 |
| pain_onset | 0.57 | 0.54 | 0.50 | 0.52 |
| pain_frequency | 0.60 | 0.57 | 0.54 | 0.55 |
| relation_to_food | 0.64 | 0.62 | 0.58 | 0.60 |
| nausea_vomiting | 0.66 | 0.64 | 0.60 | 0.62 |
| **RADIOLOGY** |  |  |  |  |
| presence_of_gallstone | 0.72 | 0.71 | 0.69 | 0.70 |
| presence_of_gallbladder | 0.93 | 0.92 | 0.92 | 0.92 |
| number_of_stones | 0.55 | 0.51 | 0.47 | 0.49 |
| diameter_of_largest_stone | 0.52 | 0.49 | 0.45 | 0.47 |
| location_of_stone | 0.57 | 0.55 | 0.52 | 0.54 |
| presence_of_gallbladder_sludge | 0.55 | 0.52 | 0.48 | 0.50 |
| cholecystitis | 0.58 | 0.56 | 0.52 | 0.54 |
| intrahepatic_duct_dilation | 0.56 | 0.54 | 0.50 | 0.52 |
| extrahepatic_duct_dilation | 0.57 | 0.55 | 0.51 | 0.53 |

